# Mapping cortical Alberta Stroke Program Early CT Score (ASPECTS) regions to functional brain networks

**DOI:** 10.1101/2025.06.20.25330017

**Authors:** Benedikt Sundermann, Christian Mathys

**Author notes:** Corresponding author: PD Dr. med. Benedikt Sundermann, Evangelisches Krankenhaus Oldenburg, Steinweg 13-17, 29129 Oldenburg, Germany.

## Abstract

This atlas-based network correspondence analysis assessed how ASPECTS regions routinely assessed in clinical acute stroke imaging are related to functional brain networks. Cortical ASPECTS regions exhibit patterns of predominant overlap with functional brain networks, albeit with limited correspondence of ASPECTS region boundaries and network boundaries. Examples are associations of the left M1 mainly with the language and fronto-parietal cognitive control networks and the M2 regions with an auditory network.

## Introduction

The Alberta Stroke Program Early CT Score (ASPECTS)^1^ is a mainstay for visually assessing the extent of acute middle cerebral artery (MCA) infarction, both in routine clinical care as well as acute stroke research. It comprises 10 regions per hemisphere (7 cortical and 3 subcortical regions). Differential contributions of infarctions in different ASPECTS regions to clinical outcomes have been reported. An example is the association of infarctions in the caudate nucleus, dorsolateral frontal (M4) and insula regions with poorer outcomes after thrombectomy regarding the modified Rankin scale at 90 days^2^.

The concept of functional brain networks is well established in basic, cognitive and clinical neuroscience. It is particularly based on functional magnetic resonance imaging research. Despite overall agreement about this concept, consensus regarding the exact spatial extent, potential overlap and nomenclature of these functional network is still evolving. A range of empirical atlases describe these networks in a 3D reference space^3^. There is increasing interest in investigating the effects of infarcts in the respective functional networks on differentiated stroke consequences such as cognitive or affective deficits. This is usually addressed in terms of stroke lesion-network-mapping^4^.

The analysis reported here addresses the knowledge gap as to how ASPECTS regions routinely assessed in clinical acute stroke imaging are related to functional brain networks.

## Methods

While not originally defined with distinct boundaries in a 3D standard space^1^, Mak et al. later provided manual masks of ASPECTS regions to be used for automated stroke imaging analyses^5^. We resampled these masks to the FSLMNI2mm space and calculated Dice coefficients representing the overlap of all 14 cortical ASPECTS region masks (insular cortex and regions M1 through M6, separately for each hemisphere) with 23 atlases (also including variants of the same basic atlases) using the network correspondence toolbox^3^ (version 0.3.3). These established atlases represent brain parcellations with predominantly cortical labels for functional brain networks. Given the lack of subcortical regions in most of these atlases, we omitted additional analyses of subcortical ASPECTS regions. Permutation tests were used to assess statistical significance. The code used for this analysis is available at https://doi.org/10.5281/zenodo.15708298.

## Results

Figure 1 shows the overlap of the cortical ASPECTS regions with the Schaefer atlas^6^ (400 parcels) with network labels by Kong et al^7^. Full results (23 atlases) are available at https://doi.org/10.5281/zenodo.15708359. Visually, there was only a relatively week correspondence of ASPECTS region boundaries and functional network boundaries. However, DICE coefficients revealed predominant and mostly symmetrical overlaps across atlases and functional network: The insular cortex ASPECTS region overlapped predominantly with the salience / ventral attention or closely related cingulo-opercular networks. The anterior ganglionic-level region (M1) correspondend with the language and fronto-parietal cognitive control networks, also exhibiting some overlap with the salience / ventral attention network. The middle middle ganglionic-level region (M2) overlapped strongly with auditory regions (as far as defined separately in the underlying atlas, particularly strong effect potentially attributatable to relatively small auditory network definition without large regions outside the middle cerebral artery territoriy). The posterior ganglionic-level region (M3) overlapped with higher visual areas and the dorsal attention network (comparatively weak and inconsistent associations across atlases). The anterior supra-ganglionic region (M4) corresponded predominantly with the fronto-parietal cognitive control network (comparatively weak and incosistent across atlases). The middle supra-ganglionic region (M5) including the pre- and postcentral gyri overlapped mainly with the somatomotor and dorsal attention networks. The posterior supra-ganglionic region (M6) overlapped with the fronto-parietal cognitive control network, default mode and dorsal attention networks, however inconsistent and relatively weak across the different atlases.

**Figure 1.**
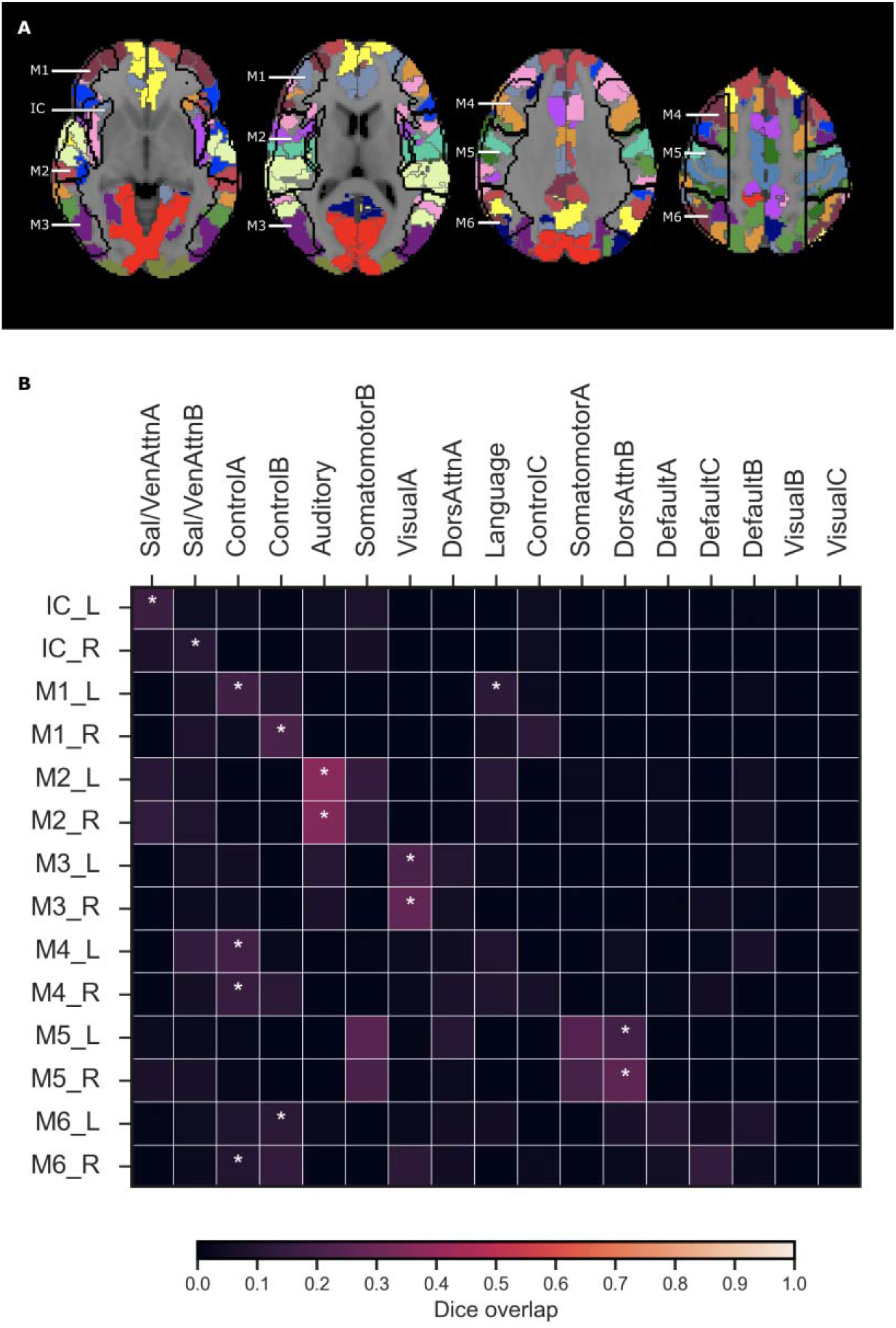
Overlap of cortical ASPECTS regions with the Schaefer atlas with region labels for 17 functional brain networks according to Kong et al. (A) Exemplary slices showing cortical ASPECTS region insular cortex (IC) and further ganglionic-level (M1-3) as well as supraganglionic (M4-6) regions as black outlines and the 17 functional networks (colors) overlaid on a brain template in the MNI standard space. (B) Dice coefficients representing the similarity of the ASPECTS regions with each network, * statistically significant overlap (p < 0.05).

## Conclusions

Cortical ASPECTS regions exhibit specific patterns of predominant correspondence with functional brain networks. Thus, assessment of individual ASPECTS regions could help leverage some information about network associations of acute MCA infarctions. Particularly the overlap with cognition-related networks was, however, relatively inconsistent and the ASPECTS regions did not align well with functional network boundaries. Given the more complex network geometry, there does not appear to be a straightforward way of just revising the definition of ASPECTS regions and their boundaries to better reflect functional networks. Thus, more detailed network-based mapping solutions are encouraged for outcome prediction based on acute stroke neuroimaging, e.g. those based on automated analysis software.

## Article Information

### Data availability

This is a secondary analysis based on published brain atlases. Materials used and results obtained in this work are referenced in the manuscript. This article does not report primary data analyses.

## References

1. Barber PA, Demchuk AM, Zhang J, Buchan AM. Validity and reliability of a quantitative computed tomography score in predicting outcome of hyperacute stroke before thrombolytic therapy. ASPECTS Study Group. Alberta Stroke Programme Early CT Score. Lancet. 2000;355:1670–1674. doi: 10.1016/s0140-6736(00)02237-6

2. Seyedsaadat SM, Neuhaus AA, Nicholson PJ, Polley EC, Hilditch CA, Mihal DC, Krings T, Benson JC, Mark I, Kallmes DF, et al. Differential Contribution of ASPECTS Regions to Clinical Outcome after Thrombectomy for Acute Ischemic Stroke. AJNR Am J Neuroradiol. 2021;42:1104–1108. doi: 10.3174/ajnr.A7096

3. Kong R, Spreng RN, Xue A, Betzel RF, Cohen JR, Damoiseaux JS, De Brigard F, Eickhoff SB, Fornito A, Gratton C, et al. A network correspondence toolbox for quantitative evaluation of novel neuroimaging results. Nat Commun. 2025;16:2930. doi: 10.1038/s41467-025-58176-9

4. Ríos AS, Temuulen U, Khalil A, Villringer K, Ali HF, Akdeniz A, Grittner U, Becher M, Rackoll T, Nave AH, et al. Lesion-Network Mapping of Post-Stroke Depressive Symptoms: Evidence from Two Prospective Ischemic Stroke Cohorts. medRxiv. 2025:2024.2012.2031.24319837. doi: 10.1101/2024.12.31.24319837

5. Mak A, Matouk CC, Avery EW, Behland J, Haider SP, Frey D, Madai VI, Vajkoczy P, Griessenauer CJ, Zand R, et al. Automated detection of early signs of irreversible ischemic change on CTA source images in patients with large vessel occlusion. PLoS One. 2024;19:e0304962. doi: 10.1371/journal.pone.0304962

6. Schaefer A, Kong R, Gordon EM, Laumann TO, Zuo XN, Holmes AJ, Eickhoff SB, Yeo BTT. Local-Global Parcellation of the Human Cerebral Cortex from Intrinsic Functional Connectivity MRI. Cereb Cortex. 2018;28:3095–3114. doi: 10.1093/cercor/bhx179

7. Kong R, Yang Q, Gordon E, Xue A, Yan X, Orban C, Zuo XN, Spreng N, Ge T, Holmes A, et al. Individual-Specific Areal-Level Parcellations Improve Functional Connectivity Prediction of Behavior. Cereb Cortex. 2021;31:4477–4500. doi: 10.1093/cercor/bhab101

